# Sars-CoV-2 attack rate in reception and accommodation centres for asylum seekers: systematic review of outbreak media reports in Germany

**DOI:** 10.1101/2021.03.12.21249641

**Authors:** Rosa Jahn, Maren Hintermeier, Kayvan Bozorgmehr

**Affiliations:** Section for Health Equity Studies and Migration, Department of General Practice and Health Services Research, University Hospital Heidelberg, Im Neuenheimer Feld 130.3, 69120 Heidelberg, Germany; Department of Population Medicine and Health Services Research, School of Public Health, Bielefeld University, D-33501 Bielefeld, Germany

**Keywords:** Sars-Cov-2, migration, refugee camps, asylum seekers, web-based systematic review

## Abstract

**Objectives:** Despite concerns about the impact of the severe acute respiratory syndrome corona virus (Sars-CoV-2) in refugee camps, data on attack rates and effectiveness of containment measures are lacking. We aimed to (1) quantify the attack rate of Sars-CoV-2 during outbreaks in reception and accommodation centres in Germany, (2) assess differences in the attack rate based on containment measures, and (3) provide an overview of testing strategies, communication, conflicts, and protection measures for refugees with special needs.

**Methods:** Systematic web-based review of outbreak media reports (until June 2020) on confirmed Sars-CoV-2 cases in reception centers for asylum seekers in Germany using the google search engine. Reports were screened for pre-defined inclusion criteria and complemented by snowball searches. Data on facility name, location, confirmed cases, containment measures, communication, protection strategies, and conflicts was extracted for each outbreak and reporting date. Evidence synthesis: meta-analysis and negative binomial regression.

**Findings:** We identified 337 media reports on 101 Sars-CoV-2 outbreaks in 99 reception and accommodation centers in Germany. The pooled Sars-CoV-2 attack rate was 13.1% (95% confidence interval, CI: 9.8-16.7). Outbreak sites implementing mass quarantine (n=76) showed higher rates (15.7; 95% CI: 11.6 - 20.2) compared to sites using conventional strategies (6.6; 95%CI: 3.1 - 11.2), yielding a rate ratio of 0.44 (95%CI: 0.27-0.72) adjusted for testing strategies, type and size of accommodation. Conflicts occurred in at least 11.8% of all outbreaks. Few sites reported specific measures to protect refugees with special needs.

**Conclusion:** Mass quarantine is associated with higher attack rates, and appears to be a counter-productive containment measure in overcrowded camps. Although further research with individual-level data is required to rule out residual confounding, reception centers and refugee camps should follow the available guidelines on Covid-19 response and refrain from mass quarantine if physical distancing cannot be guaranteed.

## 1. Introduction

Experts and international organisations from across the fields of migration, health, and human rights have highlighted the potential devastating impact of the severe acute respiratory syndrome corona virus (SARS-CoV-2) on refugee camps and expressed concern that overcrowded living conditions, limited access to health services and poor sanitation would provide fertile ground for disease transmission [1-7]. So far, confirmed cases have been reported in refugee camps in Bangladesh, Jordan, Lebanon, Syria, Greece and Palestine, and immigrant detention centres in the US [8-12]. In Germany, the first Sars-CoV-2 cases in reception centres for refugees and asylum seekers were reported in March 2020, and there have been major outbreaks with several hundred cases [13]. Asylum seekers in Germany are obliged to live in reception centres for up to 18 months with shared rooms, sanitary and kitchen facilities. As a part of the response to Sars-CoV-2 outbreaks, reception centres have repeatedly been placed under mass quarantine [14].

Despite global concerns about the transmission of Sars-CoV-2 in refugee camps and reception centres, pre-existing weaknesses in health information systems [15] lead to a lack of reliable data on attack rates as well as effectiveness and consequences of containment measures. In the absence of reliable and timely data sources to study Sars-CoV-2 outbreaks in reception centres, the extensive media coverage of such outbreaks in Germany, a country with a strong and independent press, may provide useful insights. We use a systematic web-based review strategy to (1) quantify the attack rate of Sars-CoV-2 during outbreaks among refugees living in reception and accommodation centres in Germany, (2) assess differences in the attack rate based on containment measures, and (3) provide an overview of reported testing strategies, communication, conflicts in the facilities, and protection measures for refugees with special needs.

## 2. Methods

We conducted a web-based systematic search of media reports of Sars-CoV-2 outbreaks in reception and accommodation centres in Germany, published between January 27^th^, 2020, (date of first confirmed Sars-CoV-2 case in Germany) and June 24^th^ 2020 using the Google Search Engine (see Appendix A for search queries). All retrieved hits were de-duplicated and titles and full-texts were screened for pre-defined inclusion criteria: formal media source (i.e. no social media); mention Sars-CoV-2 among refugees in Germany in title; reporting number of confirmed cases in full-text. Both search and screening were conducted in duplicate (RJ, MH) and disagreements resolved by consensus.

Media reports were clustered for each outbreak and complemented by outbreak-specific snowball searches. For each outbreak, data was extracted on: reporting date, facility name and location, incident and cumulative cases of SARS-CoV-2 infections among refugees or staff, testing strategies, quarantine measures, measures to isolate infected individuals, conflicts within the facility as well as communication strategies. Outbreaks were excluded if the total number of inhabitants of the centre (at-risk population) was not reported.

The attack rate was calculated as the cumulative number of confirmed cases per outbreak divided by the at-risk population. In case of disagreement between reported numbers of inhabitants of a centre, the mean was used as denominator. Attack rates were pooled (i) across all outbreak sites, (ii) by strictest form of management strategy applied over the course of the outbreak (mass-quarantine vs. no mass-quarantine) and (iii) by accommodation type (reception centers (RC); district accommodation centres (AC)) using random effects models with the Freeman and Tukey double arcsine transformation. Meta-analysis was performed using the ‘metaprop’ command in StataSE 15 [16]. Funnelplots were used as graphical test of bias (using Stata’s ‘metafunnel’ command) [17]. We performed two sensitivity analyses to account for: 1) potential superspreading events by excluding outbreak-sites with more than 50% of inhabitants infected, and 2) potential effects of testing strategies on attack rates by stratified analyses (mass testing vs. targeted testing of contact persons or symptomatic individuals). We further analysed the relationship between outbreak management strategies and Sars-CoV-2 attack rate by multiple regression in a negative binomial model (‘nbreg’ command), controlling for possible confounders at level of facilities (testing strategy, and size and type of accommodation centres).

## 3. Results

The search strategy yielded a total of 337 reports which were included for analysis (see table 1 for PRISMA flow chart). Of these, 196 (58,16%) were published by local newspapers, 45 (13,35%) by regional and national newspaper outlets, and 44 (13,06%) were press releases by local governments or city administrations. Other sources included web reports from radio (n=30; 8,90%) and TV stations (n=7; 2,08%), and political outlets (n=9; 2,67%). The median number of reports per outbreak was 3.71 (min:1; max:15) and for 91 outbreaks (90,10%), at least two reports were available.

**Table 1:** PCR-confirmed SARS-CoV-2 cases per outbreak and population size per centre, overall and by centre type, N=101 outbreaks in 14 federal states, Germany.

|  | Reception Centre (RC) |  | Accommodation Centre (AC) |  | Overall |  |
| --- | --- | --- | --- | --- | --- | --- |
|  | Cases (n) | Inhabitants (N) | Cases (n) | Inhabitants (N) | Cases (n) | Inhabitants (N) |
| **Mean** | 55.77 | 360.27 | 15.95 | 121.16 | 26.2 | 182.71 |
| **SD** | 86.35 | 215.47 | 19.72 | 125.17 | 49.58 | 185.04 |
| **Md** | 20.5 | 309.5 | 7 | 88 | 8 | 118 |
| **Min** | 1 | 39 | 1 | 11 | 1 | 11 |
| **Max** | 400 | 792 | 86 | 850 | 400 | 850 |

*Legend: n= PCR-confirmed SARS-CoV-2 cases, N= Total number of inhabitants (at-risk population) SD= standard deviation; Md= Median; Min=Minimum; Max=Maximum*

### 3.1 Descriptive analysis of review results

We identified 101 COVID-19 outbreaks in 99 reception and accommodation centers across 14 of the 16 German federal states. 2,646 confirmed SARS-CoV-2 infections were reported among a total of 18,454 residents, as well as 81 confirmed cases among staff. 26 of these outbreaks occurred in RC and 75 in AC (see table 1).

Mass quarantine, i.e. indiscriminate movement restriction of all inhabitants and restriction of in-and-out movements, was implemented during 76 outbreaks (75% of all outbreaks), affecting a total of 12,692 refugees. The average duration of mass quarantine was 19 days, with a high variation (SD: 8.62 min. 2; max. 43). Of all sites implementing mass quarantine (N=76), 84.2% (n=64) implemented this measure within two days after the first confirmed SARS-CoV-2 case. In 23.8% (n=24) of all outbreaks (N=101), conventional management strategies were applied, i.e. isolation of confirmed cases with or without contact tracing and quarantine of close contact persons.

Efforts to isolate confirmed cases from the remaining inhabitants were reported in 64 (84.21%) of the 76 outbreak sites which were placed under mass quarantine. Among sites applying conventional management strategies (N=24) or where this information was missing (n=1), the isolation of infected individuals was reported for 23 outbreaks (92%). Specific measures to protect individuals with special needs, commonly comprising unaccompanied minors, elderly individuals, or pregnant women, were reported for 27 (26.7%) outbreaks. Of these, 17 (63.0%) sites evacuated or transferred refugees with special needs to separate areas within the center or to designated protective shelters.

Across the 101 identified outbreaks, mass testing of all inhabitants of the centre was implemented in 75.3% (n=61), with some centres repeating the mass testing every few days. Among the centres implementing mass quarantine (N=76), 65,8% (n=50) implemented mass testing at least once. In 11.9% (n=12) of all outbreaks (N=101), tests were conducted for contact persons of confirmed cases, and 7.9% (n=8) of outbreak sites followed other strategies, such as testing individuals with clinical symptoms only. Information on testing strategies was not reported for 19.8% (n=20) of outbreaks.

Specific measures to inform the centres’ inhabitants about the Covid-19 pandemic or specific containment measures were reported in 25.7% (n=26) of outbreaks, 9.0% (n=9) reported that no specific measures were taken, the remainder lacked data on these aspects. In 11.8% (n=12) of all outbreaks, conflicts were reported within the facilities. These occurred mostly in connection with mass quarantine measures and often necessitated police response. In 10.8% (n=11) it was explicitly stated there had been no conflicts, while reports on the remaining outbreaks lacked information on conflicts.

### 3.2 Pooled Sars-CoV-2 attack rate

The pooled Sars-CoV-2 attack rate for the 101 outbreaks in accommodation centers for asylum seekers was 13.08% (95%CI: 9.84-16.69), and no differences were observed between different accommodation types (RC: 12.93% (95%CI:6.39-21.28), AC: 13.11% (95%CI:9.88-16.70)). Attack rates were higher among outbreak sites under mass quarantine (15.65% (95%CI:11.58-20.18)) compared to outbreaks in which conventional management strategies were applied (6.60% (95%CI:3.09-11.17)) (see figure 2).

**Figure 1:**
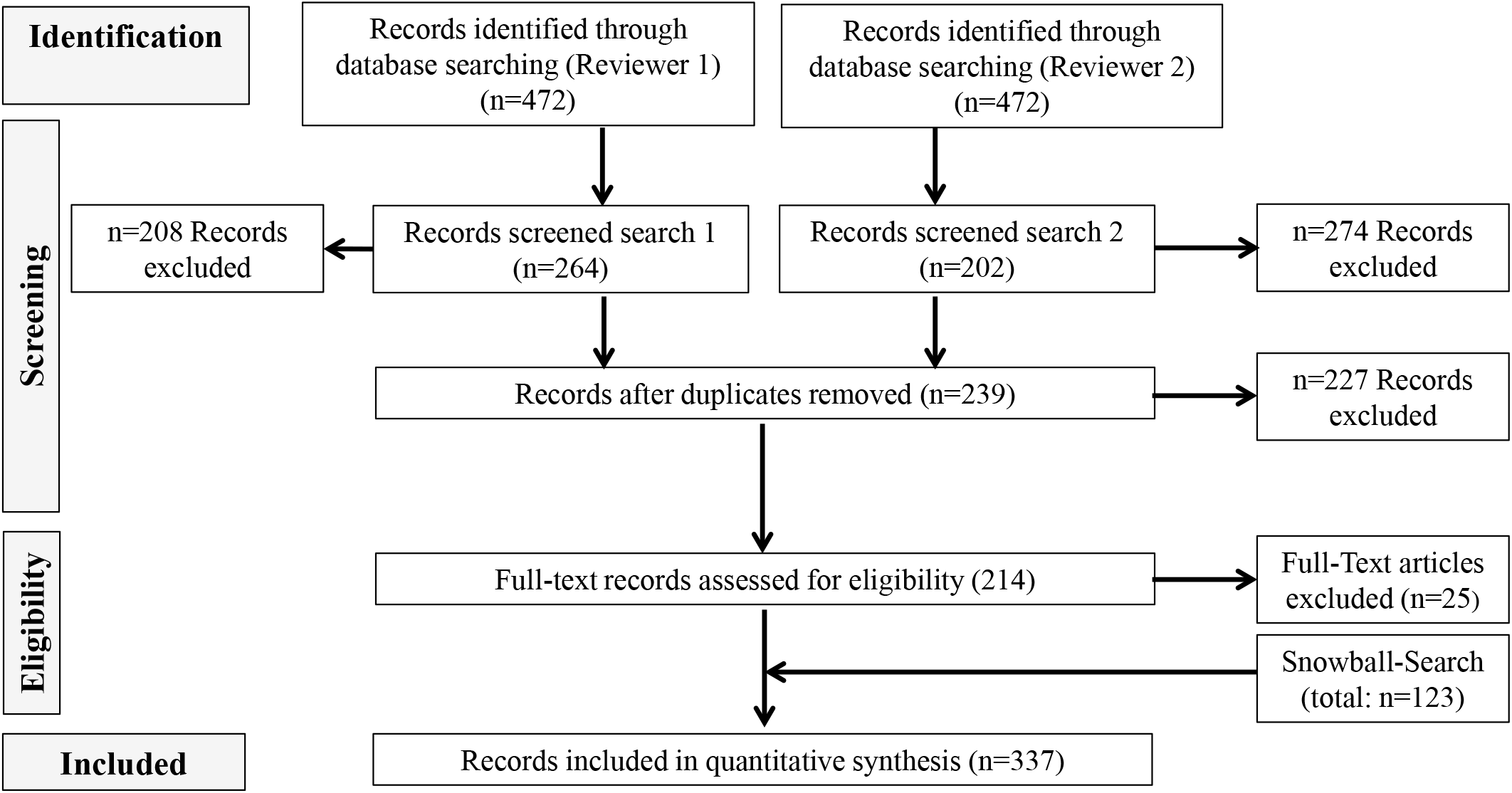
PRISMA flow chart.

**Figure 2:**
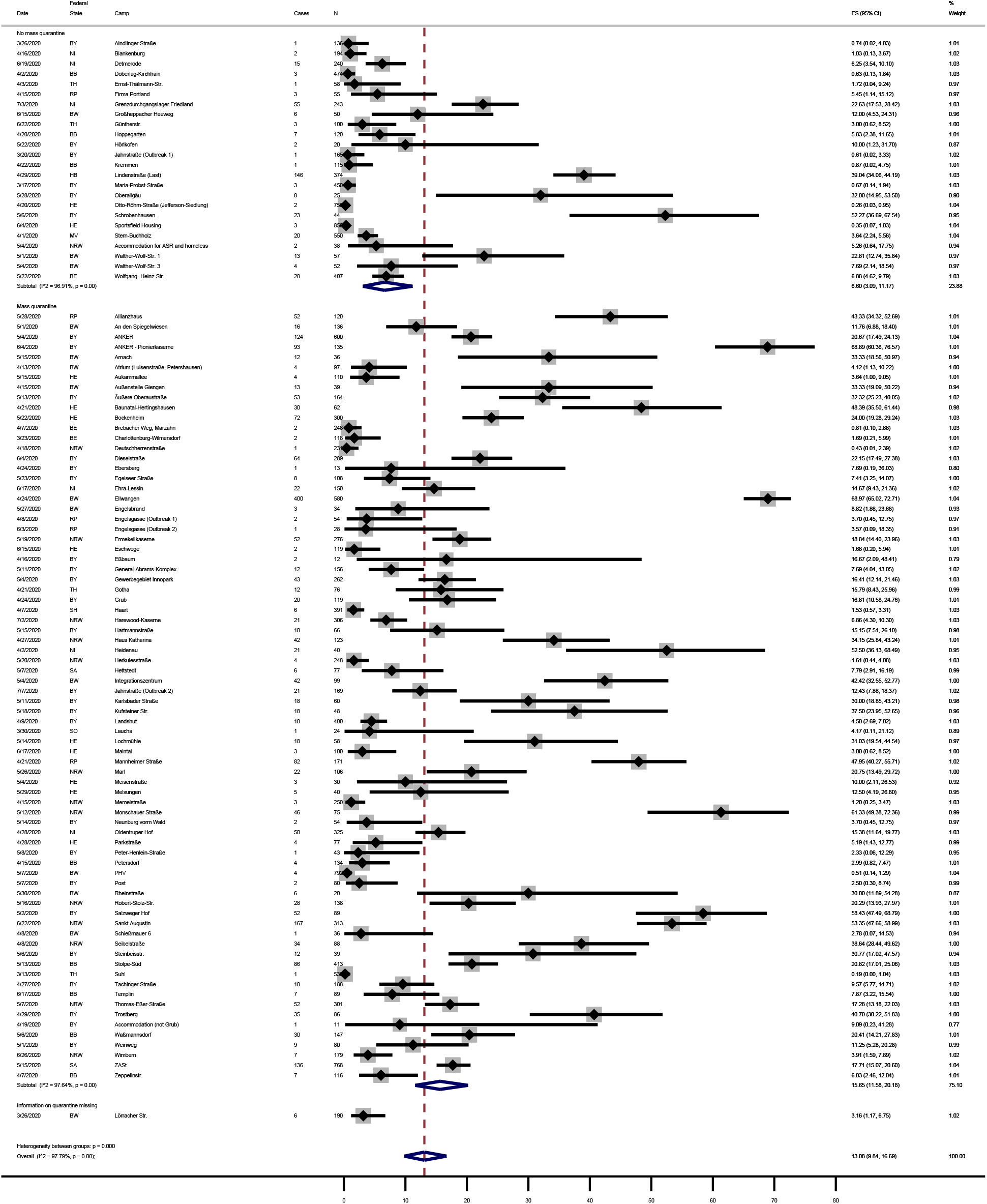
Forest plot of attack rates for each facility and pooled estimates, by type of quarantine, N=101 outbreaks in 14 federal states, Germany. Legend: ES: estimate of attack rates. I^2^: I-squared measure of heterogeneity. Information on quarantine measures was not reported for one outbreak (missing = 1). Y-axis: attack rate in percentages. Federal states: BB: Brandenburg; BE: Berlin; BW: Baden-Württemberg; BY: Bavaria; HB: Bremen; HE: Hesse; MV: Mecklenburg Western Pomerania; NI: Lower Saxony; NW: Northrhine-Westphalia; RP: Rhineland Palatinate; SN: Saxony; ST: Saxony-Anhalt; SH: Schleswig Holstein; TH: Thuringia.

The funnel plot showed an asymmetric distribution, and grouping by quintiles of inhabitants shows a tendency towards lower attack rates in larger camps, likely due to a higher number of non-contact persons considered as “at-risk population”. Stratified analysis by accommodation size showed no difference in attack rates (Appendix B). The egger’s test rejects the H0-hypothesis of no small-study effect (p= 0.000), indicating that a small-study effect may be a possible explanation for asymmetric distribution (Appendix C).

Excluding seven outbreak sites that could represent super-spreading events (attack rates > 50%) (sensitivity analysis 1) reduced the overall pooled attack rate to 10.05% (95%CI: 8.05; 13.21), but did not affect the finding that attack rates in facilities under mass quarantine were higher (12.59%; 95%CI: 9.63 - 15.87) compared to sites applying conventional management strategies (5.38%; (95%CI: 2.39 - 9.34) (for details see Appendix D). Sensitivity analysis 2 revealed higher Sars-CoV-2 attack rates in outbreaks implementing mass testing (16.05%; 95%CI: 11.38 - 21.32) compared to sites implementing targeted testing of close contacts or only symptomatic inhabitants (9.07%; 95%CI: 4.12 - 15.57) (for details see Appendix D). Testing strategies differed by quarantine measures to marginally significant (p=0.053) extent (table 2).

**Table 2:** Absolute and relative frequency of outbreaks by quarantine measure and testing strategy.

| Quarantine measure | Mass testing<br>n (%) | Targeted testing or test-information N/A*<br>n (%) | Total<br>N (%) |
| --- | --- | --- | --- |
| Mass quarantine | 50 (65.79%) | 26 (34.21%) | 76 (100%) |
| No mass quarantine + | 11 (44.00%) | 14 (56.00%) | 25 (100%) |
| Total | 61 (60.40%) | 40 (39.60%) | 101 (100%) |

Pearson chi2(1) = 3.7340 p = 0.053
Legend: Pearson chi2: chi-square test statistic. p: p-value. \*Information on testing strategy not available (not reported). +Includes n=1 centre for which Information on quarantine measures was missing/not reported. n: absolute frequency. %: percentage. N= totals.

The attack rate among outbreaks implementing conventional containment strategies was 0.44 times the rate under mass quarantine, adjusted for testing strategy, as well as size and type of accommodation (table 3).

**Table 3:** Rate ratios of SARS-CoV-2 attack rates (per 100.000) obtained by multiple negative binomial regression model, N=100 outbreaks in 14 federal states, Germany.

|  | Variable (vs. reference) | RR [95% CI] | p-value |
| --- | --- | --- | --- |
| **Quarantine measure** | No mass quarantine<sup>+</sup><br>(vs. mass quarantine) | 0.44 [0.27-0.72] | 0.001 |
| **Testing strategy** | Targeted testing or test-information N/A<br>(vs. mass testing) | 0.63 [0.41-0.97] | 0.034 |
| **Accommodation size** | Q2 | 0.96 [0.50-1.84] | 0.904 |
|  | Q3 | 0.56 [0.29-1.09] | 0.089 |
|  | Q4 | 0.49 [0.26-0.96] | 0.038 |
|  | Q5 | 0.56 [0.26-1.18] | 0.128 |
|  | (vs. Q1) |  |  |
| **Accommodation type** | Reception centre<br>(vs. district accommodation centre) | 1.38 [0.76-2.50] | 0.284 |

*Legend: RR: rate ratio. N/A: no information available. vs: versus. Q1-5: quintiles. <sup>+</sup>Includes n=1 centre for which information on quarantine measures was missing/not reported.*

## 4. Discussion

Using a web-based systematic review approach, we found a Sars-CoV-2 attack rate of 13% in reception and accommodation centers for asylum seekers in Germany. Outbreak management strategies included mass quarantine of entire centers among 75% of the 101 identified outbreaks. In these settings, the Sars-CoV-2 attack rate was significantly higher compared to conventional management strategies. The difference in Sars-CoV-2 attack rates between sites implementing mass quarantine and those using conventional strategies remained stable when excluding outbreaks with potential super-spreading events (sensitivity analysis 1), and when controlling in multiple regression models for testing strategies as well as accommodation type and size. Information on conflicts was rare, but they occurred in at least about 12% of all outbreaks. Few sites reported specific measures for the protection of refugees with special needs.

Our findings conform with other studies in this context. The Robert Koch Institute reported 199 outbreaks in German reception and accommodation centers for asylum seekers recorded by the national notification system by August 2020 [18]. The outbreaks comprised in average 20.8 confirmed Sars-CoV-2 cases, the highest average outbreak size among all reported outbreak locations in Germany [18]. However, no attack rates can be calculated based on data of the national notification system as it contains data on cases only and lacks data on denominators (i.e. the number of inhabitants in the centres). A modelling study of Sars-CoV-2 in the Rohingya refugee camp in Cox’s Bazar, Bangladesh found that one single case in the camp would likely lead to a large-scale outbreak with more than 1,000 cases, due to large household sizes as well as inadequate access to sanitation and hygiene [19]. Other institutionalized settings, such as prisons or nursing homes have been similarly affected by Sars-CoV-2 and overcrowding of the facilities in conjunction with a particularly vulnerable population are considered key factors in the spread of the disease. A modelling study on outbreaks in elderly homes in Ontario, Canada, found that a reduction of individuals per room from four to two could have prevented 19% of all infections, and 18% of all deaths [20]. Reflecting such findings, European and international guidelines on Covid-19 containment measures in refugee camps and other institutionalized settings recommend the reduction of inhabitants to allow for physical distancing and self-isolation, isolation of confirmed cases and quarantine of contact persons only, and infectious disease surveillance [2, 21-23].

Our review shows that mass quarantine is used as a rule, rather than an exception of outbreak management strategies in reception centers for refugees in Germany, despite not being recommended by the available Covid-19 guidelines for refugee camps. The finding of higher attack rates in centers under mass quarantine compared to conventional approaches supports concerns raised by academia [24], the Robert Koch Institute [23], the ECDC (13) and civil society organisations that mass quarantine may increase transmission risk within facilities due to lack of possibilities to self-isolate and perform social distancing. Comparisons can be drawn between the situation of reception centers under mass quarantine and outbreaks of Covid-19 in closed setting such as the cruise ship Diamond princess. Here, the cumulative incidence risk was found to be 17%, and it is worth noting that several national governments, including the US and Germany, considered it to be high enough to warrant the evacuation of their citizens from the ship [25, 26]. Modelling studies suggest asymptomatic patients (64%) contributed to the spread of the disease on board, and that measures to separate infected individuals as well as reducing contact between passengers had lowered the basis reproduction rate over the course of the outbreak [27].

However, our results show that even while under mass quarantine, measures to reduce disease transmission within the facilities were not sufficiently adopted. For example, 34.2% of facilities under mass quarantine were reported not to conduct series tests to identify asymptomatic cases, and 15.8% reportedly did not strictly separate infected from non-infected individuals. Mass quarantine of reception centers therefore may not only increase the risk of conflicts, stigma, or mental health disorders [28, 29], but it is also associated with higher risk of transmission in camp contexts and does not offer adequate protection for vulnerable individuals. This finding is consistent with modelling studies suggesting early evacuation is more effective to reduce transmission in confined contexts such as cruise ships [30] compared to mass quarantine. While more studies on management strategies are needed, the study findings suggest that mass quarantine should not only be avoided for ethical or psychosocial reasons, but also on epidemiological grounds.

### 4.1. Strengths and limitations

The results of this study are the first estimate of the attack rate of Sars-Cov-2 infection in refugee reception centers that we are aware of, especially as no such studies were found in available systematic reviews on Covid-19 in migrants [31]. The reason for a lack of studies lies in weak health information systems that do not have the capacity to generate timely and reliable health data in reception centers and refugee camps [15]. Given the lack of timely and reliable data, the web-based systematic review approach has proven to be a useful tool to generate early estimates while retaining an acceptable degree of robustness. A similar approach has been applied by Dawood et al. (2020), using daily web-based surveillance to identify global patterns of Sars-CoV-2 infections [32]. The sources they included focused on official government or ministry websites. While we searched for official press releases for the identified outbreaks of Covid-19 in reception centers, these constituted 13% of the included reports. We did, however, cross-match information from available reports on each of the 91 (90,10%) outbreaks for which more than one report was available to verify the reported data. Limitations associated with the web-based systematic review approach include incompleteness of some of the contextual data regarding testing strategies, information of inhabitants, conflicts, and protection measures for at-risk individuals. Information on quarantine measures and dates on which measures have been implemented, however, was available for all but one of the outbreaks. There may be an over reporting of more severe outbreaks, possibly because mass quarantine measures or police presence in the camps may attract attention from the host community as well as the media, leading to ascertainment bias. Moreover, differing testing strategies and time lags in the testing of inhabitants may have resulted in a delayed diagnosis and the infections may have occurred before the quarantine measures took effect. However, as most facilities implemented mass quarantine within two days after the first SARS-CoV-2 case was confirmed, the measure was likely to precede diagnosis of further cases, so that reverse causation (i.e., higher incidence leading to more strict containment strategies such as mass quarantine) can be considered unlikely. To better understand the dynamic of these local outbreaks and to rule out residual confounding, further research using individual-level data in respective outbreak sites is urgently needed. Moreover, the significant heterogeneity between centers suggest relevant contextual factors in disease transmission which warrant more detailed study.

## 5. Conclusion

The estimates from this web-based systematic review show Sars-Cov-2 attack rates of 13% in German reception centers. Attack rates were higher under mass quarantine compared to conventional strategies, even after controlling for differences in testing strategies and other facility-level variables. This suggests that mass quarantine does not benefit overall virus containment in camps which do not allow for self-isolation and physical distancing, and may hence be counter-productive. Although further research with individual-level data is required to rule out residual confounding, authorities should refrain from implementing mass quarantine in reception centers and refugee camps if physical distancing cannot be guaranteed and follow conventional strategies (eventually complemented by mass testing). Implementation of the available guidelines on prevention of Covid-19 in refugee camps, aiming at reducing the number of individuals per accommodation unit, providing each household with individual sanitary facilities, and promoting access to hygiene and personal protective equipment such as face masks, is paramount to effective prevention and control of Sars-CoV-2.

## Data Availability

All data / links to media reports can pe provided upon request.

## 6. Ethical issues

The research did not involve human subjects and used anonymous media reports. No ethical clearance was required.

