## Appendix A for "Sars-CoV-2 attack rate in reception and accommodation centres for asylum seekers: systematic review of outbreak media reports in Germany": Appendix_A_Search queries.docx

**Appendix 1: Search queries**

Search queries were developed based on a preliminary web search for reports on Sars-CoV-2 outbreaks in reception and accommodation centers for asylum seekers in Germany. Using the terms most commonly used to describe such outbreaks, we developed four search queries. These consisted of the term “refugee”, key words for “reception centers” (facility, reception center, accommodation center, asylum home/center) and “corona infection” or “corona cases” (see table 1).

***Table 1: Search queries (German and English translation)***

| **Search query** | **Searchterm (German)** | **Translation** |
| --- | --- | --- |
| Search 1 | Einrichtung Flüchtlinge Corona Infektion | Facility refugees corona infection |
| Search 2 | Erstaufnahme ANKER Flüchtlinge Corona Fälle | Reception center refugees corona cases |
| Search 3 | Gemeinschaftsunterkunft Flüchtlinge Corona Fälle | Accommodation center refugees corona cases |
| Search 4 | Unterkunft Heim Asyl Corona Fälle | Center asylum home corona cases |
