## Appendix B for "Sars-CoV-2 attack rate in reception and accommodation centres for asylum seekers: systematic review of outbreak media reports in Germany"

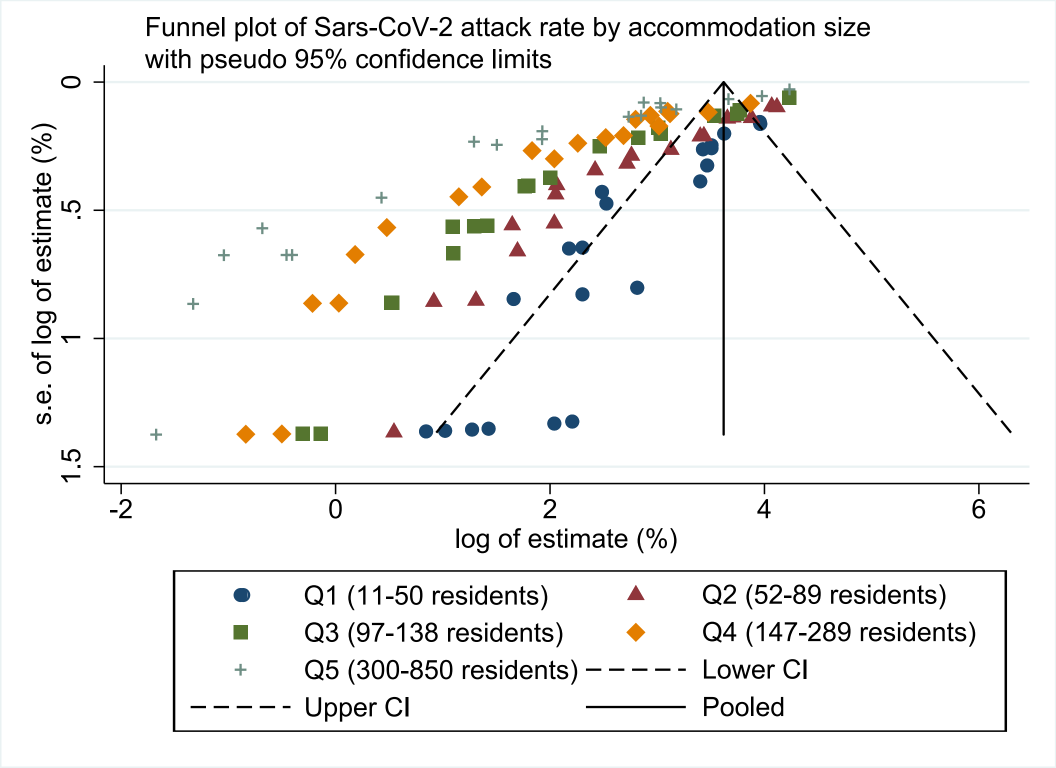
Funnel plot of Sars-CoV-2 attack rate by accommodation size
with pseudo 95% confidence limits
s.e. of log of estimate (%)
0
.5
1
1.5
-2
0
2
4
6
log of estimate (%)
Q1 (11-50 residents)
Q2 (52-89 residents)
Q3 (97-138 residents)
Q4 (147-289 residents)
Q5 (300-850 residents)
Lower CI
Upper CI
Pooled

### Forest plot of Sars-CoV-2 attack rate for each facility and pooled estimates, by accommdation size

| Date | Federal State | Camp | Cases | N | ES (95% CI) | % Weight |
|---|---|---|---|---|---|---|
| Q3 (97-138 residents) | | | | | | |
| 3/26/2020 | BY | Aindlinger Straße | 1 | 136 | 0.74 (0.02, 4.03) | 1.01 |
| 5/28/2020 | RP | Allianzhaus | 52 | 120 | 43.33 (34.32, 52.69) | 1.01 |
| 5/1/2020 | BW | An den Spiegelwiesen | 16 | 136 | 11.76 (6.88, 18.40) | 1.01 |
| 6/4/2020 | BY | ANKER - Pionierkaserne | 93 | 135 | 68.89 (60.36, 76.57) | 1.01 |
| 4/13/2020 | BW | Atrium (Luisenstraße, Petershausen) | 4 | 97 | 4.12 (1.13, 10.22) | 1.00 |
| 5/15/2020 | HE | Aukammallee | 4 | 110 | 3.64 (1.00, 9.05) | 1.01 |
| 3/23/2020 | BE | Charlottenburg-Wilmersdorf | 2 | 118 | 1.69 (0.21, 5.99) | 1.01 |
| 5/23/2020 | BY | Egelseer Straße | 8 | 108 | 7.41 (3.25, 14.07) | 1.00 |
| 6/15/2020 | HE | Eschwege | 2 | 119 | 1.68 (0.20, 5.94) | 1.01 |
| 4/24/2020 | BY | Grub | 20 | 119 | 16.81 (10.58, 24.76) | 1.01 |
| 6/22/2020 | TH | Güntherstr. | 3 | 100 | 3.00 (0.62, 8.52) | 1.00 |
| 4/27/2020 | NRW | Haus Katharina | 42 | 123 | 34.15 (25.84, 43.24) | 1.01 |
| 4/20/2020 | BB | Hoppegarten | 7 | 120 | 5.83 (2.38, 11.65) | 1.01 |
| 5/4/2020 | BW | Integrationszentrum | 42 | 99 | 42.42 (32.55, 52.77) | 1.00 |
| 4/22/2020 | BB | Kremmen | 1 | 115 | 0.87 (0.02, 4.75) | 1.01 |
| 6/17/2020 | HE | Maintal | 3 | 100 | 3.00 (0.62, 8.52) | 1.00 |
| 5/26/2020 | NRW | Marl | 22 | 106 | 20.75 (13.49, 29.72) | 1.00 |
| 4/15/2020 | BB | Petersdorf | 4 | 134 | 2.99 (0.82, 7.47) | 1.01 |
| 5/16/2020 | NRW | Robert-Stolz-Str. | 28 | 138 | 20.29 (13.93, 27.97) | 1.01 |
| 4/7/2020 | BB | Zeppelinstr. | 7 | 116 | 6.03 (2.46, 12.04) | 1.01 |
| Subtotal (I^2 = 96.54%, p = 0.00) | | | | | 11.53 (5.42, 19.46) | 20.15 |
| Q5 (300-850 residents) | | | | | | |
| 5/4/2020 | BY | ANKER | 124 | 600 | 20.67 (17.49, 24.13) | 1.04 |
| 5/22/2020 | HE | Bockenheim | 72 | 300 | 24.00 (19.28, 29.24) | 1.03 |
| 4/2/2020 | BB | Doberlug-Kirchhain | 3 | 474 | 0.63 (0.13, 1.84) | 1.03 |
| 4/24/2020 | BW | Ellwangen | 400 | 580 | 68.97 (65.02, 72.71) | 1.04 |
| 4/7/2020 | SH | Haart | 6 | 391 | 1.53 (0.57, 3.31) | 1.03 |
| 7/2/2020 | NRW | Harewood-Kaserne | 21 | 306 | 6.86 (4.30, 10.30) | 1.03 |
| 4/9/2020 | BY | Landshut | 18 | 400 | 4.50 (2.69, 7.02) | 1.03 |
| 4/29/2020 | HB | Lindenstraße (Last) | 146 | 374 | 39.04 (34.06, 44.19) | 1.03 |
| 3/17/2020 | BY | Maria-Probst-Straße | 3 | 450 | 0.67 (0.14, 1.94) | 1.03 |
| 4/28/2020 | NI | Oldentruper Hof | 50 | 325 | 15.38 (11.64, 19.77) | 1.03 |
| 4/20/2020 | HE | Otto-Röhm-Straße (Jefferson-Siedlung) | 2 | 75[illegible] | 0.26 (0.03, 0.95) | 1.04 |
| 5/7/2020 | BW | PHV | 4 | 792 | 0.51 (0.14, 1.29) | 1.04 |
| 6/22/2020 | NRW | Sankt Augustin | 167 | 313 | 53.35 (47.66, 58.99) | 1.03 |
| 6/4/2020 | HE | Sportsfield Housing | 3 | 85[illegible] | 0.35 (0.07, 1.03) | 1.04 |
| 4/1/2020 | MV | Stern-Buchholz | 20 | 550 | 3.64 (2.24, 5.56) | 1.04 |
| 5/13/2020 | BB | Stolpe-Süd | 86 | 413 | 20.82 (17.01, 25.06) | 1.03 |
| 3/13/2020 | TH | Suhl | 1 | 53[illegible] | 0.19 (0.00, 1.04) | 1.03 |
| 5/7/2020 | NRW | Thomas-Eßer-Straße | 52 | 301 | 17.28 (13.18, 22.03) | 1.03 |
| 5/22/2020 | BE | Wolfgang- Heinz-Str. | 28 | 407 | 6.88 (4.62, 9.79) | 1.03 |
| 5/15/2020 | SA | ZASt | 136 | 768 | 17.71 (15.07, 20.60) | 1.04 |
| Subtotal (I^2 = 99.32%, p = 0.00) | | | | | 10.71 (4.44, 19.23) | 20.67 |
| Q1 (11-50 residents) | | | | | | |
| 5/15/2020 | BW | Arnach | 12 | 36 | 33.33 (18.56, 50.97) | 0.94 |
| 4/15/2020 | BW | Außenstelle Giengen | 13 | 39 | 33.33 (19.09, 50.22) | 0.94 |
| 4/24/2020 | BY | Ebersberg | 1 | 13 | 7.69 (0.19, 36.03) | 0.80 |
| 5/27/2020 | BW | Engelsbrand | 3 | 34 | 8.82 (1.86, 23.68) | 0.93 |
| 6/3/2020 | RP | Engelsgasse (Outbreak 2) | 1 | 28 | 3.57 (0.09, 18.35) | 0.91 |
| 4/16/2020 | BY | Eßbaum | 2 | 12 | 16.67 (2.09, 48.41) | 0.79 |
| 6/15/2020 | BW | Großheppacher Heuweg | 6 | 50 | 12.00 (4.53, 24.31) | 0.96 |
| 4/2/2020 | NI | Heidenau | 21 | 40 | 52.50 (36.13, 68.49) | 0.95 |
| 5/22/2020 | BY | Hörlkofen | 2 | 20 | 10.00 (1.23, 31.70) | 0.87 |
| 5/18/2020 | BY | Kufsteiner Str. | 18 | 48 | 37.50 (23.95, 52.65) | 0.96 |
| 3/30/2020 | SO | Laucha | 1 | 24 | 4.17 (0.11, 21.12) | 0.89 |
| 5/4/2020 | HE | Meisenstraße | 3 | 30 | 10.00 (2.11, 26.53) | 0.92 |
| 5/29/2020 | HE | Melsungen | 5 | 40 | 12.50 (4.19, 26.80) | 0.95 |
| 5/28/2020 | BY | Oberallgäu | 8 | 25 | 32.00 (14.95, 53.50) | 0.90 |
| 5/8/2020 | BY | Peter-Henlein-Straße | 1 | 43 | 2.33 (0.06, 12.29) | 0.95 |
| 5/30/2020 | BW | Rheinstraße | 6 | 20 | 30.00 (11.89, 54.28) | 0.87 |
| 4/8/2020 | BW | Schießmauer 6 | 1 | 36 | 2.78 (0.07, 14.53) | 0.94 |
| 5/6/2020 | BY | Schrobenhausen | 23 | 44 | 52.27 (36.69, 67.54) | 0.95 |
| 5/6/2020 | BY | Steinbeisstr. | 12 | 39 | 30.77 (17.02, 47.57) | 0.94 |
| 4/19/2020 | BY | Accommodation (not Grub) | 1 | 11 | 9.09 (0.23, 41.28) | 0.77 |
| 5/4/2020 | NRW | Accommodation for ASR and homeless | 2 | 38 | 5.26 (0.64, 17.75) | 0.94 |
| Subtotal (I^2 = 82.41%, p = 0.00) | | | | | 17.52 (10.76, 25.36) | 19.08 |
| Q4 (147-289 residents) | | | | | | |
| 5/13/2020 | BY | Äußere Oberaustraße | 53 | 164 | 32.32 (25.23, 40.05) | 1.02 |
| 4/16/2020 | NI | Blankenburg | 2 | 194 | 1.03 (0.13, 3.67) | 1.02 |
| 4/7/2020 | BE | Brebacher Weg, Marzahn | 2 | 248 | 0.81 (0.10, 2.88) | 1.03 |
| 6/19/2020 | NI | Detmerode | 15 | 240 | 6.25 (3.54, 10.10) | 1.03 |
| 4/18/2020 | NRW | Deutschherrenstraße | 1 | 231 | 0.43 (0.01, 2.39) | 1.02 |
| 6/4/2020 | BY | Dieselstraße | 64 | 289 | 22.15 (17.49, 27.38) | 1.03 |
| 6/17/2020 | NI | Ehra-Lessin | 22 | 150 | 14.67 (9.43, 21.36) | 1.02 |
| 5/19/2020 | NRW | Ermekeilkaserne | 52 | 276 | 18.84 (14.40, 23.96) | 1.03 |
| 5/11/2020 | BY | General-Abrams-Komplex | 12 | 156 | 7.69 (4.04, 13.05) | 1.02 |
| 5/4/2020 | BY | Gewerbegebiet Innopark | 43 | 262 | 16.41 (12.14, 21.46) | 1.03 |
| 7/3/2020 | NI | Grenzdurchgangslager Friedland | 55 | 243 | 22.63 (17.53, 28.42) | 1.03 |
| 5/20/2020 | NRW | Herkulesstraße | 4 | 248 | 1.61 (0.44, 4.08) | 1.03 |
| 3/20/2020 | BY | Jahnstraße (Outbreak 1) | 1 | 165 | 0.61 (0.02, 3.33) | 1.02 |
| 7/7/2020 | BY | Jahnstraße (Outbreak 2) | 21 | 169 | 12.43 (7.86, 18.37) | 1.02 |
| 3/26/2020 | BW | Lörracher Str. | 6 | 190 | 3.16 (1.17, 6.75) | 1.02 |
| 4/21/2020 | RP | Mannheimer Straße | 82 | 171 | 47.95 (40.27, 55.71) | 1.02 |
| 4/15/2020 | NRW | Memelstraße | 3 | 250 | 1.20 (0.25, 3.47) | 1.03 |
| 4/27/2020 | BY | Tachinger Straße | 18 | 188 | 9.57 (5.77, 14.71) | 1.02 |
| 5/6/2020 | BB | Waßmannsdorf | 30 | 147 | 20.41 (14.21, 27.83) | 1.01 |
| 6/26/2020 | NRW | Wimbern | 7 | 179 | 3.91 (1.59, 7.89) | 1.02 |
| Subtotal (I^2 = 96.68%, p = 0.00) | | | | | 9.53 (5.14, 15.04) | 20.44 |
| Q2 (52-89 residents) | | | | | | |
| 4/21/2020 | HE | Baunatal-Hertingshausen | 30 | 62 | 48.39 (35.50, 61.44) | 0.98 |
| 4/8/2020 | RP | Engelsgasse (Outbreak 1) | 2 | 54 | 3.70 (0.45, 12.75) | 0.97 |
| 4/3/2020 | TH | Ernst-Thälmann-Str. | 1 | 58 | 1.72 (0.04, 9.24) | 0.97 |
| 4/15/2020 | RP | Firma Portland | 3 | 55 | 5.45 (1.14, 15.12) | 0.97 |
| 4/21/2020 | TH | Gotha | 12 | 76 | 15.79 (8.43, 25.96) | 0.99 |
| 5/15/2020 | BY | Hartmannstraße | 10 | 66 | 15.15 (7.51, 26.10) | 0.98 |
| 5/7/2020 | SA | Hettstedt | 6 | 77 | 7.79 (2.91, 16.19) | 0.99 |
| 5/11/2020 | BY | Karlsbader Straße | 18 | 60 | 30.00 (18.85, 43.21) | 0.98 |
| 5/14/2020 | HE | Lochmühle | 18 | 58 | 31.03 (19.54, 44.54) | 0.97 |
| 5/12/2020 | NRW | Monschauer Straße | 46 | 75 | 61.33 (49.38, 72.36) | 0.99 |
| 5/14/2020 | BY | Neunburg vorm Wald | 2 | 54 | 3.70 (0.45, 12.75) | 0.97 |
| 4/28/2020 | HE | Parkstraße | 4 | 77 | 5.19 (1.43, 12.77) | 0.99 |
| 5/7/2020 | BY | Post | 2 | 80 | 2.50 (0.30, 8.74) | 0.99 |
| 5/2/2020 | BY | Salzweger Hof | 52 | 89 | 58.43 (47.49, 68.79) | 1.00 |
| 4/8/2020 | NRW | Seibelstraße | 34 | 88 | 38.64 (28.44, 49.62) | 1.00 |
| 6/17/2020 | BB | Templin | 7 | 89 | 7.87 (3.22, 15.54) | 1.00 |
| 4/29/2020 | BY | Trostberg | 35 | 86 | 40.70 (30.22, 51.83) | 1.00 |
| 5/1/2020 | BW | Walther-Wolf-Str. 1 | 13 | 57 | 22.81 (12.74, 35.84) | 0.97 |
| 5/4/2020 | BW | Walther-Wolf-Str. 3 | 4 | 52 | 7.69 (2.14, 18.54) | 0.97 |
| 5/1/2020 | BY | Weinweg | 9 | 80 | 11.25 (5.28, 20.28) | 0.99 |
| Subtotal (I^2 = 93.92%, p = 0.00) | | | | | 18.10 (10.49, 27.14) | 19.67 |
| Heterogeneity between groups: p = 0.164 | | | | | | |
| Overall (I^2 = 97.79%, p = 0.00); | | | | | 13.08 (9.84, 16.69) | 100.00 |

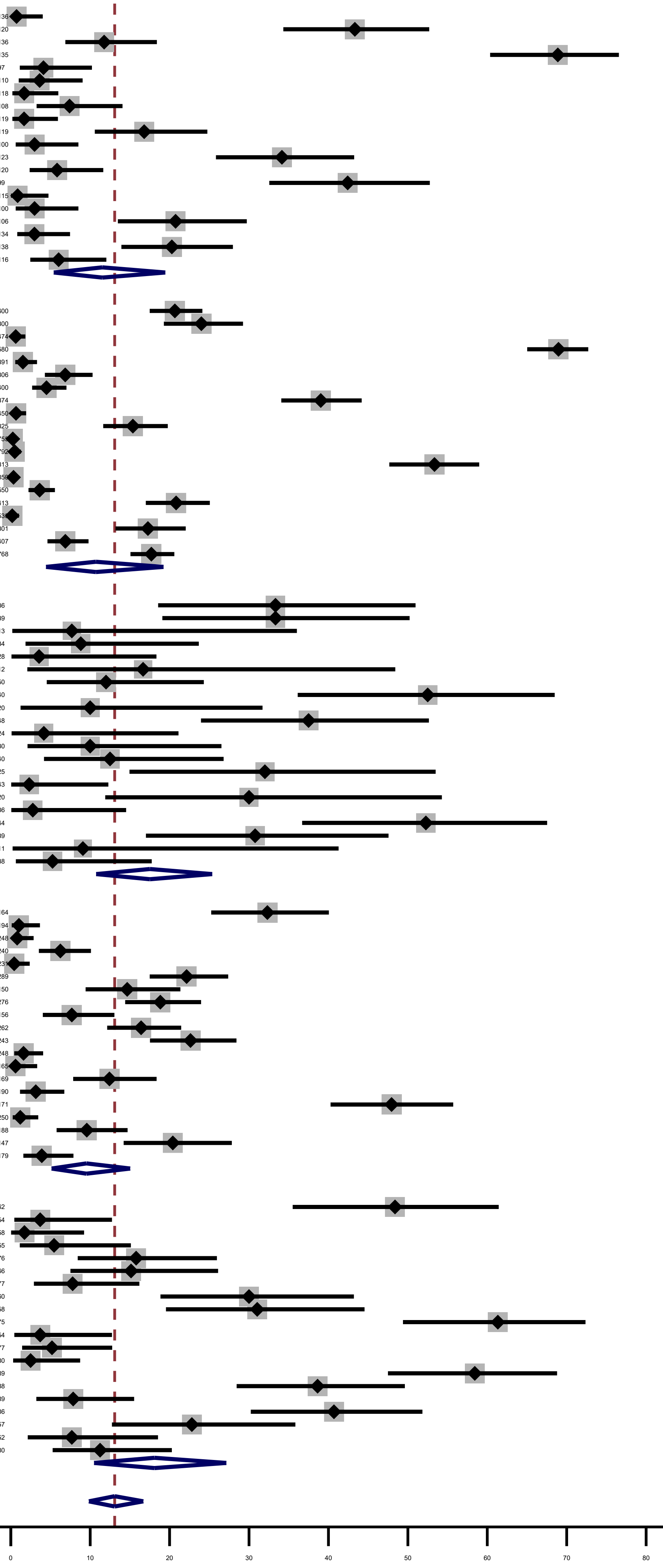
