## Supplementary figures and images for "Sars-CoV-2 attack rate in reception and accommodation centres for asylum seekers: systematic review of outbreak media reports in Germany"

### Appendix C

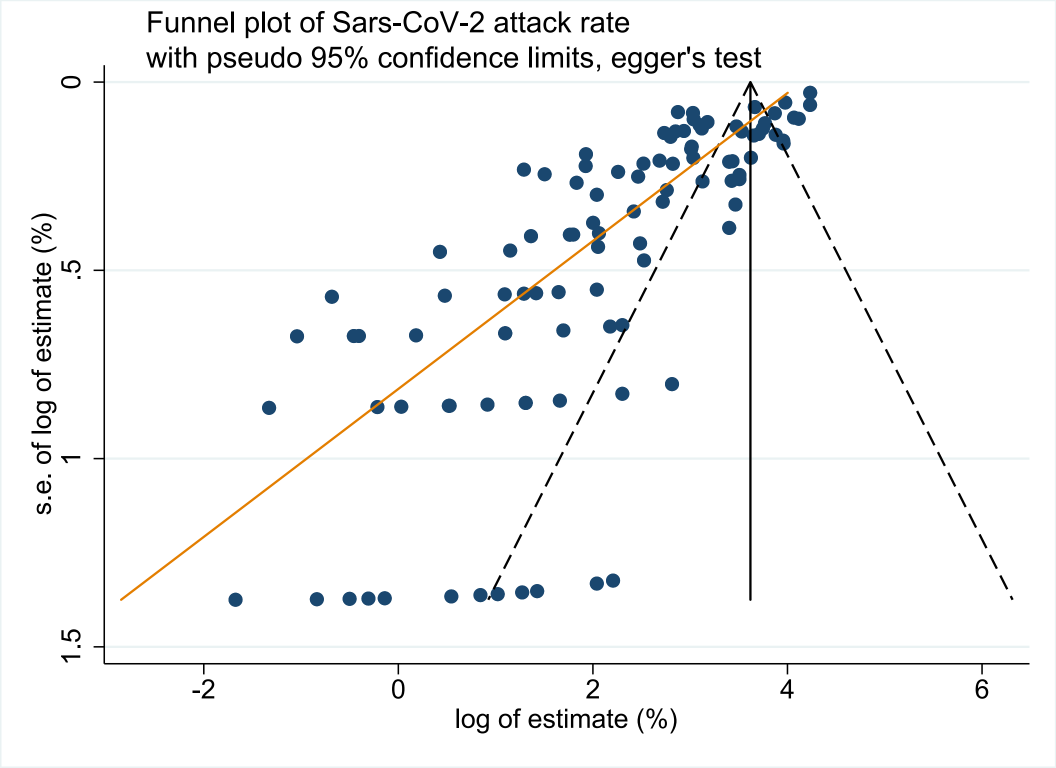
